# The basic reproduction number of COVID-19 across Africa

**DOI:** 10.1101/2021.11.02.21265826

**Authors:** Sarafa A. Iyaniwura, Musa Rabiu, Jummy F. David, Jude D. Kong

## Abstract

The pandemic of the severe acute respiratory syndrome coronavirus 2 (SARS-CoV-2) took the world by surprise. Following the first outbreak of COVID-19 in December 2019, several models have been developed to study and understand its transmission dynamics. Although the spread of COVID-19 is being slowed down by vaccination and other interventions, there is still a need to have a clear understanding of the evolution of the pandemic across countries, states and communities. To this end, there is a need to have a clearer picture of the initial spread of the disease in different regions. In this project, we used a simple SEIR model and a Bayesian inference framework to estimate the basic reproduction number of COVID-19 across Africa. Our estimates vary between 1.98 (Sudan) and 9.66 (Mauritius), with a median of 3.67 (90% CrI: 3.31 - 4.12). The estimates provided in this paper will help to inform COVID-19 modeling in the respective countries/regions.

## 1 Introduction

The COVID-19 pandemic started with an outbreak of the disease in the city of Wuhan, Hubei province, China, in December 2019 [110]. COVID-19 is a respiratory infectious disease caused by the severe acute respiratory syndrome coronavirus 2 (SARS-CoV-2) [8]. It is believed to be transmitted through the inhalation of virus-infected droplets, coughing, sneezing and having physical contact with infected persons (direct transmission) or indirectly when a susceptible individual comes in contact with contaminated commonly shared surface or object [58, 25, 82, 111, 37]. Following the outbreak of the disease, several non-pharmaceutical interventions (NPIs) such as physical distancing, self-isolation, hand washing, stay-at-home order, closing of schools/businesses, travel restrictions, among others, were deployed worldwide to control its spread [100, 18, 33, 10, 35, 63, 87]. Despite implementing these NPIs, COVID-19 continued to spread all over the world causing a significant number of infections and deaths in the first few months following its outbreak. The World Health Organization (WHO) declared COVID-19 a public health emergency of international concern (PHEIC) on January 20, 2020 [112] and a pandemic on March 11, 2020 [113]. As of October 29, 2021, there had been over 245 million confirmed cases of COVID-19 and over 4.97 million deaths globally [114]. In addition to infections and deaths caused by COVID-19, the pandemic has had a significant effect on our day-to-day activities such as socializing, entertainment, education, tourism, business, health, and so on [14, 15, 81].

The first case of COVID-19 in Africa was reported in Egypt on February 14, 2020 [45, 83]. As of October 29, 2021, forty-seven (47) African countries have been affected by the disease, with over 6.07 million confirmed cases and over 150,00 confirmed deaths [47]. During the initial phase of COVID-19, Africa was regarded as one of the most vulnerable continents due to its large volume of commercial activities with China and its deplorable health system [45, 83]. As a result of this, some African countries implemented interventions such as travel restriction and border closure [45, 109, 36] even before their first case of the disease to prevent case importation from the COVID-19 epicenters (such as Spain, China, United States of America, United Kingdom etc.). The mitigation of local transmission of COVID-19 after case importation depends mainly on the timing and how strict other interventions such as physical distancing, wearing of face mask, closure of schools and businesses, prevention of public gathering, etc., are implemented. Table A1 of Appendix A shows when the first case of COVID-19 was reported in the African countries we considered and the interventions implemented during the early stages of the diseases in these countries.

To understand the transmission dynamics of COVID-19, several mathematical models have been developed and analyzed [62, 74, 88, 89, 78, 98, 55, 54, 120, 6, 104, 22, 10, 65]. Some of these models have also been used to study the effectiveness of different intervention strategies aimed at controlling the spread of the disease and to inform public health policies [10, 55, 104, 17, 5]. Understanding the early dynamics of a disease is essential to model its transmission dynamics adequately. In mathematical modeling of infectious diseases, the basic reproduction number is used to capture the initial growth rate of a disease in a completely susceptible population. The basic reproduction number (*R*_0_) of an infectious disease is the expected number of secondary cases of the disease caused directly by a single infected individual introduced into a completely susceptible population [108, 31, 107, 86]. It gives us a better understanding of how contagious the disease is and how fast it will spread in a completely susceptible population without any interventions. Following a disease outbreak in a population, it is essential to estimate its *R*_0_ to enable public health officers and government officials to plan appropriately for the disease and implement intervention strategies to prevent its spread.

Numerous models have been developed to estimate the basic reproduction number of COVID-19 across the world [85, 84, 68, 75, 119, 60, 19, 34, 61]. In [61], logistic growth curves were fitted to the daily reported cases of COVID-19 for 58 countries across the world. They estimated the basic reproduction number of COVID-19 for these countries and determined the social, economical, and environmental factors that influence the difference in this parameter across these countries. The exponential growth rate and basic reproduction number of COVID-19 for Africa were calculated in [75] using the method proposed by [68] together with the Poisson likelihood method for data fitting. They used the serial interval estimated from COVID-19 cases in Hong Kong, China [118] and estimated the exponential growth rate of COVID-19 in Africa to be 0.22 per day (95% CI: 0.20 - 0.24) and the basic reproduction number as 2.37 (95% CI: 2.22 - 2.51). The COVID-19 basic reproduction number for the 15 largest countries in Western Europe were estimated using the exponential growth rate approach in [66]. In [117], the maximum likelihood method and the sequential Bayesian method were used to calculate the COVID-19 basic reproduction number (*R*_0_) and the time-varying estimate of the effective reproduction number (*R*_*t*_), respectively, for the 12 countries that were most affect by COVID-19 in the early days of the pandemic. Other papers that estimated the basic reproduction number for COVID-19 in different regions include [30], where they estimated *R*_0_ for Sri Lanka using three different approaches: a SIR compartmental model, exponential growth rate method, and the maximum likelihood method. For the SIR model, they obtained *R*_0_ = 1.02 with confidence interval (CI) of 0.75-1.29; 0.93 (CI: 0.77–1.10) from the exponential growth rate method; and 1.23 (CI: 0.94–1.57) using the maximum likelihood method. The variation in the value of *R*_0_ they obtained using these three methods ranges from 0.69 to 2.20. In [1], the basic reproduction of COVID-19 in Nigeria was estimated by fitting an SEIR-type model to the cumulative reported cases and death data. In addition, they used a likelihood-based approach to estimate the effective reproduction number *R*(*t*) in a Bayesian framework.

Although there is a decline in the daily reported cases of COVID-19 globally, there is still a significant spread of the disease worldwide. On October 29, 2021, over 456,520 cases of COVID-19 were reported globally within 24 hours with over 8,000 confirmed deaths [114]. Due to the continuing spread of COVID-19, there is a need to adequately understand the evolution of the disease across countries, states and communities. To this end, there is a need to understand the initial dynamics of the disease in different regions across the world. In this study, we use a simple susceptible-exposed-infectious-recovered (SEIR) model together with a Bayesian inference framework to estimate the basic reproduction number (*R*_0_) for COVID-19 across Africa. These estimates are based on the reported number of COVID-19 cases in these countries during the early stage of the epidemic before non-pharmaceutical interventions aimed at mitigating community transmissions were implemented. The remaining part of this paper is organized as follows: In Section 2.1, we present the simple SEIR model used in this study. A brief discussion about the data sources is presented in Section 2.2, and in Section 2.3, we describe our Bayesian inference framework. Results and discussions are presented in Section 3 followed by a conclusion.

## 2 Methods

### 2.1 Mathematical Model

We use a simple susceptible-exposed-infectious-recovered (SEIR) compartmental model to estimate the basic reproduction number of COVID-19 across Africa. This model has four compartments namely: susceptible (*S*), exposed (*E*), infectious (*I*) and recovered/removed (*R*). Individuals in the exposed compartment do not transmit the disease [40, 65, 64], while infectious individuals in *I* can transmit infection to their contacts at some rate. Similar to the recently published articles [60, 13, 30, 76, 73, 99], our infectious compartment *I* contains both symptomatic and asymptomatic individuals. We assume that recovered individuals do not get re-infected during the epidemic period that we considered. The SEIR model considered in this paper is given by

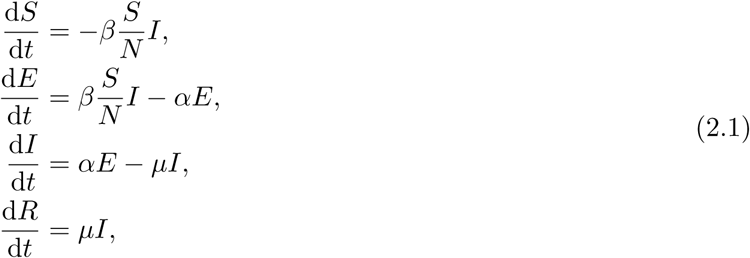

where *β* is the transmission rate, *α* is the rate of transitioning from the exposed compartment (*E*) to the infectious compartment (*I*), and *µ* is the recovery rate. It can easily be verified using the next-generation matrix approach [31, 108, 57] that the basic reproduction number for (2.1) satisfies

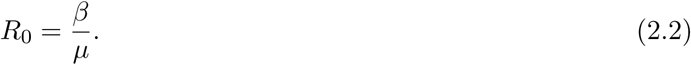

We observe from (2.2) that the basic reproduction number is directly proportional to the disease transmission rate (*β*) and inversely proportional to the recovery rate (*µ*). This implies that *R*_0_ will increase as the transmission rate increases and decrease as the recovery rate increases. The cumulative predicted cases of the model is computed from individuals transitioning from the exposed compartment (*E*) to the infectious compartment (*I*).

Table 1 shows the model variables and their description. In addition, it contains the parameters of the model, their descriptions and values. The rate of transitioning from exposed to infectious compartment (*α*) and the infectious recovery rate (*µ*) were obtained from published literature, while the transmission rate (*β*) is estimated for each country from the daily reported cases using Bayesian inference. The basic reproduction number for each country is then computed using (2.2) and the estimated transmission rate.

**Table 1:**
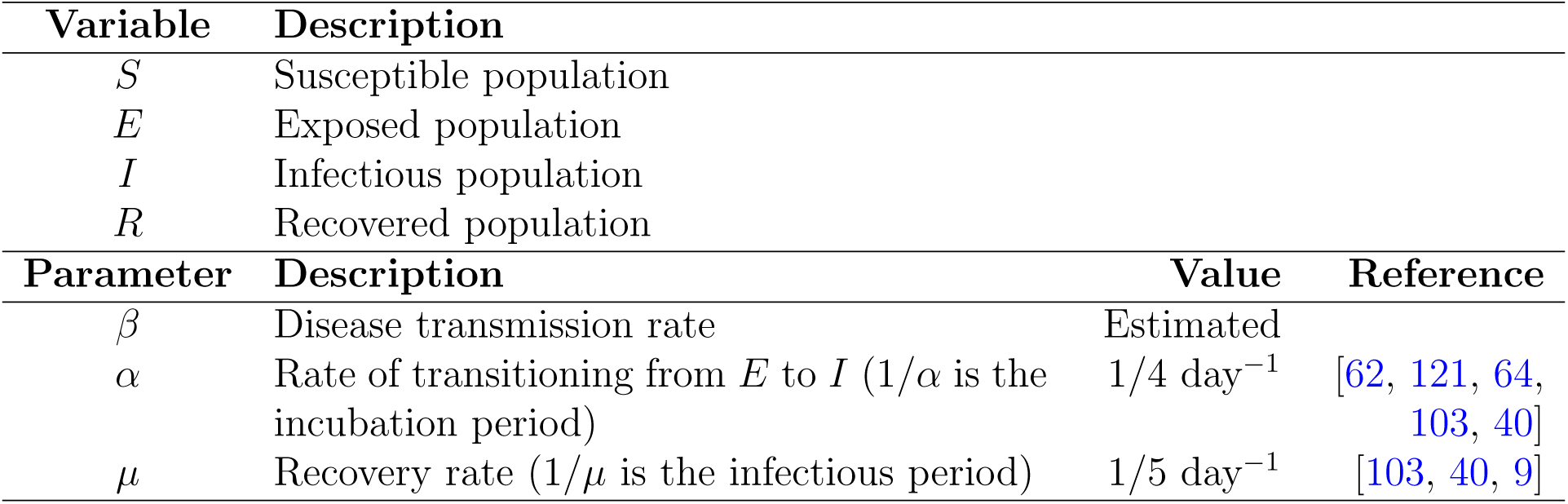
Model variables, parameters, descriptions and values.

### 2.2 Data

We collected the daily reported cases of COVID-19 for each country from the publicly available data set on the GitHub page of *Our World in Data* [53, 52]. The countries considered in this study are selected based on the availability of data. For island countries or regions, we ensure that their total population size is over 500, 000. In addition, we ensured that none of the countries considered has any major conflict or war.

We considered 24 countries and divided them into regions as follows. West African: Nigeria, Ghana, Senegal and Mali; Southern Africa: South Africa, Zambia, Namibia and Malawi; North Africa: Egypt, Tunisia, Algeria and Morocco; East Africa: Ethiopia, Kenya, Rwanda and Sudan; Central Africa: Cameroon, Chad, Gabon and Congo; and African Islands: Madagascar, Comoros, Mauritius and Cape Verde. When there is a missing data in the first five days of the epidemic, we set the case number to zero; otherwise, we use the average of the cases reported before and after the missing data. Since we are interested in estimating the basic reproduction number, only the first few days of the epidemic were considered to capture the evolution of disease at the early stage of the outbreak. Hence, the number of data points considered differ by country depending on the scenarios in each country.

Table A1 and A2 of Appendix A show the countries, their population sizes, the date of their first case of COVID-19, and the non-pharmaceutical interventions implemented during the early stages of their outbreak. We fit the predicted cases of our SEIR model (2.1) to the daily reported number of COVID-19 cases collected for each country from *Our World in Data* Github page [53] and estimate the transmission rate of the disease using Bayesian inference. Using (2.2) and the estimated transmission rate, we estimate the corresponding basic reproduction number for each country. In our SEIR model (2.1), the number of predicted daily cases is computed as the number of exposed individuals (*E*) transitioning to the infectious class (*I*).

### 2.3 Bayesian inference

The SEIR model (2.1) was fitted to the daily reported cases of COVID-19 collected for each country from the publicly available data set taken from *Our World in Data* [53], using a Bayesian inference framework and the RStan package in R version 3.6.3 [101]. The Bayesian inference framework enables us to incorporate our prior knowledge into the model parameters and also gives us the ability to evaluate probabilistic statements about the data given the model. For each country, the likelihood is constructed as follows

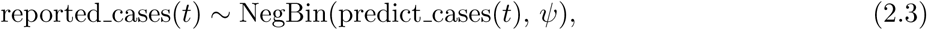

where reported cases(*t*) is the daily reported cases of COVID-19 in the country at time *t*, predict cases(*t*) is the predicted daily cases computed from the SEIR model (2.1), NegBin(·) is the negative binomial distribution function in RStan, and *ψ* is the over-dispersion parameter. To ensure that our SEIR model (2.1) is coded accurately in the Stan function and also for validation, we generated synthetic cases data using our model with known parameter values. We tested the model’s ability to recover these parameter values by fitting it to the synthesized data set. The resulting posterior distributions were inspected for biases and coverage of the true parameter values used to generate the synthetic data. All the inferences presented in this paper were done using the adaptive Hamiltonian Monte Carlo method No-U-Turn sampling (NUTS) in RStan with 5,000 iterations and 4 chains. See [49, 70] for more details about the NUTS.

## 3 Results and discussion

We fitted the simple SEIR model (2.1) to the daily reported number of COVID-19 cases in selected African countries using a Bayesian inference framework. We estimated the basic reproduction number of COVID-19 for these countries. The results of our inferences showing the daily reported cases together with the predicted cases obtained from our model are presented in Figures 1 and 2. The black dots are the daily reported cases of COVID-19 for each country, while the solid lines are the median predicted cases. The narrower (darker) bands are 50% credible intervals (CrI), while the wider (lighter) bands are 90% CrI. The estimated *R*_0_ values are plotted in Figure 3 with 90% CrI and their values given in Table 2 with 90% CrI. We have grouped the countries into regions for organizational and comparison purposes.

**Table 2:**
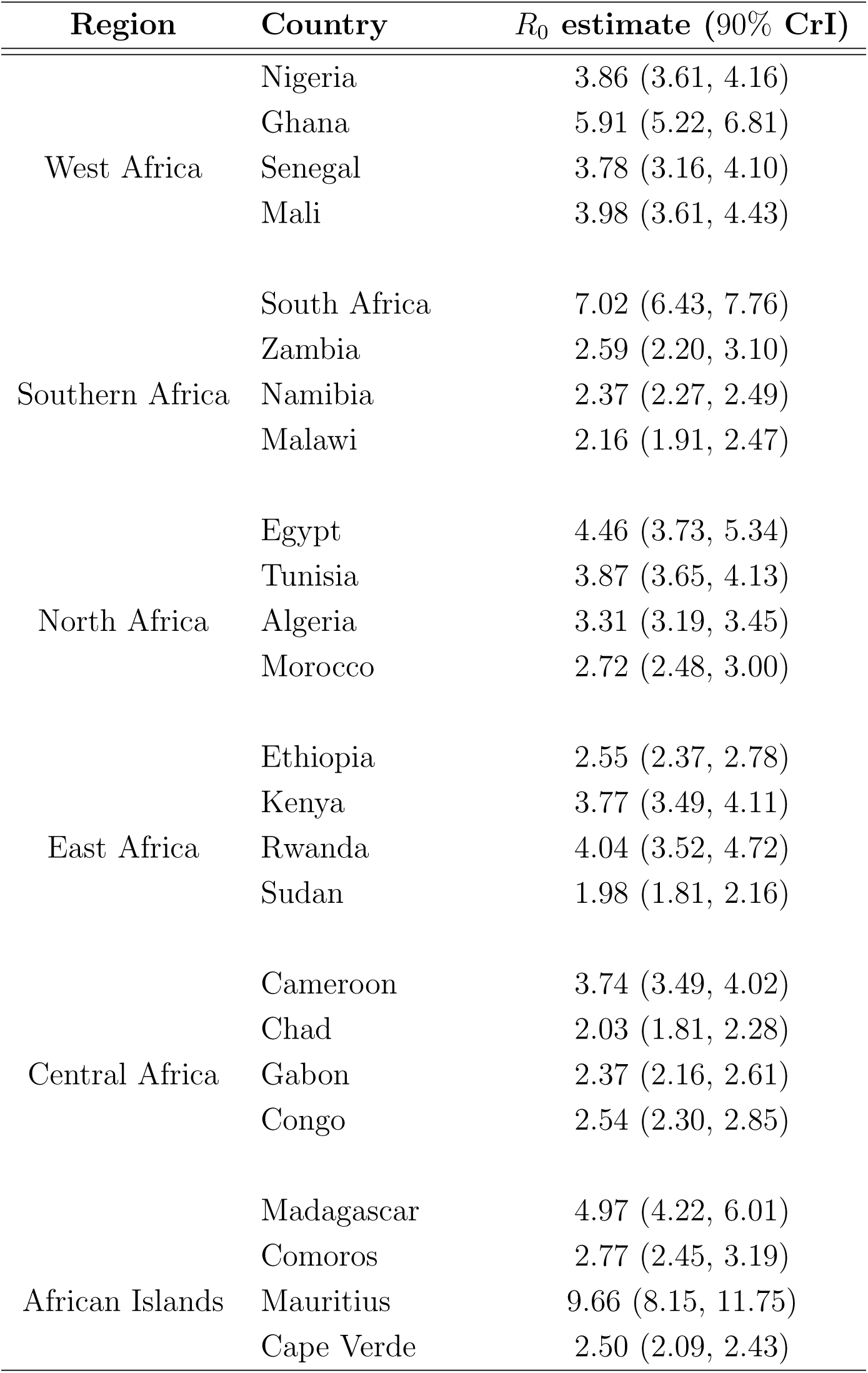
Estimated basic reproduction number. The estimated basic reproduction number for each country with 90% credible interval (CrI).

| Region | Country | $R_0$ estimate (90% CrI) |
| --- | --- | --- |
| West Africa | Nigeria | 3.86 (3.61, 4.16) |
|  | Ghana | 5.91 (5.22, 6.81) |
|  | Senegal | 3.78 (3.16, 4.10) |
|  | Mali | 3.98 (3.61, 4.43) |
| Southern Africa | South Africa | 7.02 (6.43, 7.76) |
|  | Zambia | 2.59 (2.20, 3.10) |
|  | Namibia | 2.37 (2.27, 2.49) |
|  | Malawi | 2.16 (1.91, 2.47) |
| North Africa | Egypt | 4.46 (3.73, 5.34) |
|  | Tunisia | 3.87 (3.65, 4.13) |
|  | Algeria | 3.31 (3.19, 3.45) |
|  | Morocco | 2.72 (2.48, 3.00) |
| East Africa | Ethiopia | 2.55 (2.37, 2.78) |
|  | Kenya | 3.77 (3.49, 4.11) |
|  | Rwanda | 4.04 (3.52, 4.72) |
|  | Sudan | 1.98 (1.81, 2.16) |
| Central Africa | Cameroon | 3.74 (3.49, 4.02) |
|  | Chad | 2.03 (1.81, 2.28) |
|  | Gabon | 2.37 (2.16, 2.61) |
|  | Congo | 2.54 (2.30, 2.85) |
| African Islands | Madagascar | 4.97 (4.22, 6.01) |
|  | Comoros | 2.77 (2.45, 3.19) |
|  | Mauritius | 9.66 (8.15, 11.75) |
|  | Cape Verde | 2.50 (2.09, 2.43) |

**Figure 1:**
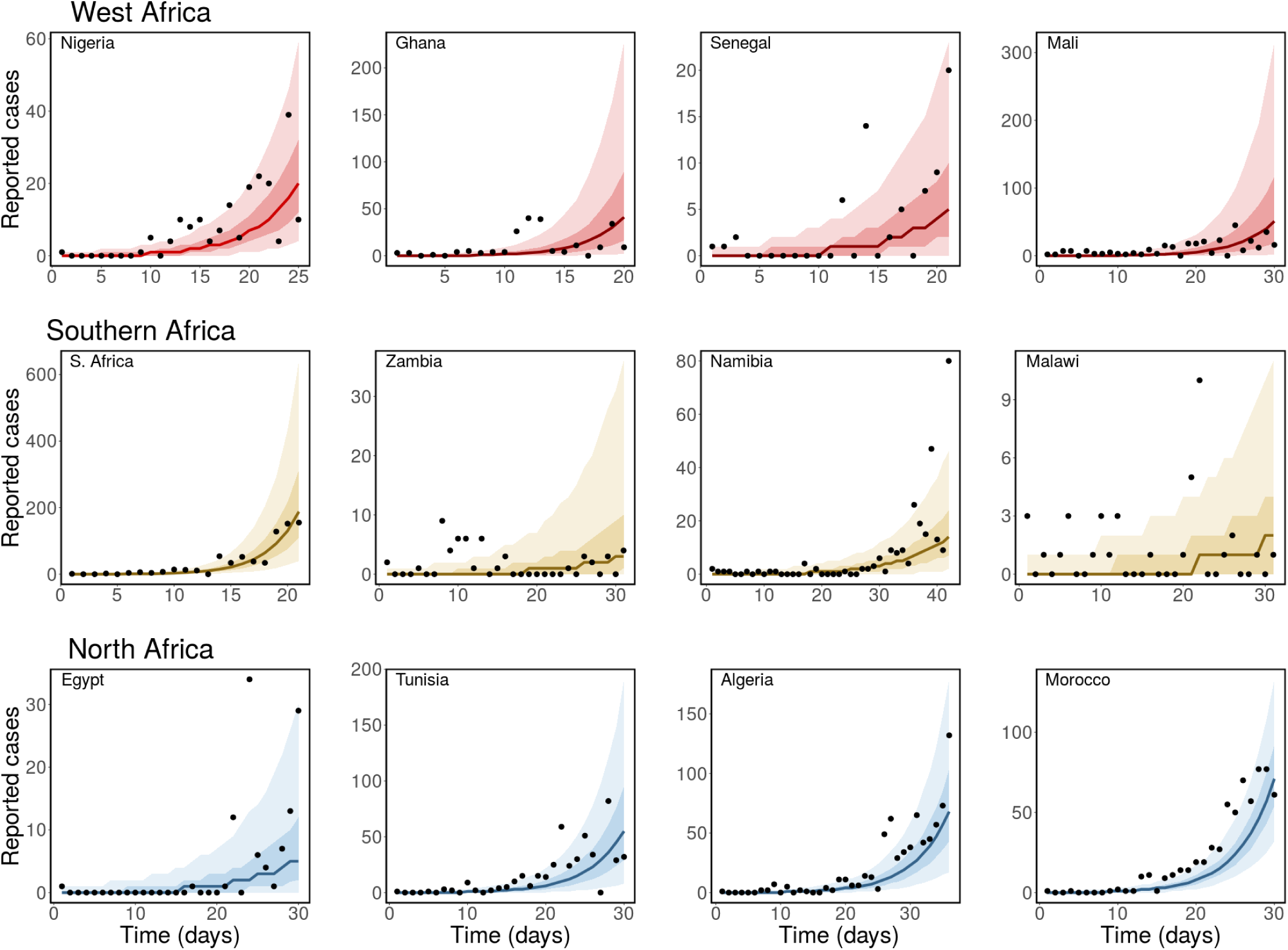
Observed and estimated number of daily cases of COVID-19. Top panel: West Africa (Nigeria, Ghana, Senegal and Mali), middle panel: Southern Africa (South Africa, Zambia, Namibia and Malawi), and bottom panel: North Africa (Egypt, Tunisia, Algeria and Morocco). The black dots are the daily reported cases of COVID-19, the solid lines are the median predicted cases, the narrow (darker) bands are the 50% CrI, while the wider (lighter) bands are the 90% CrI.

**Figure 2:**
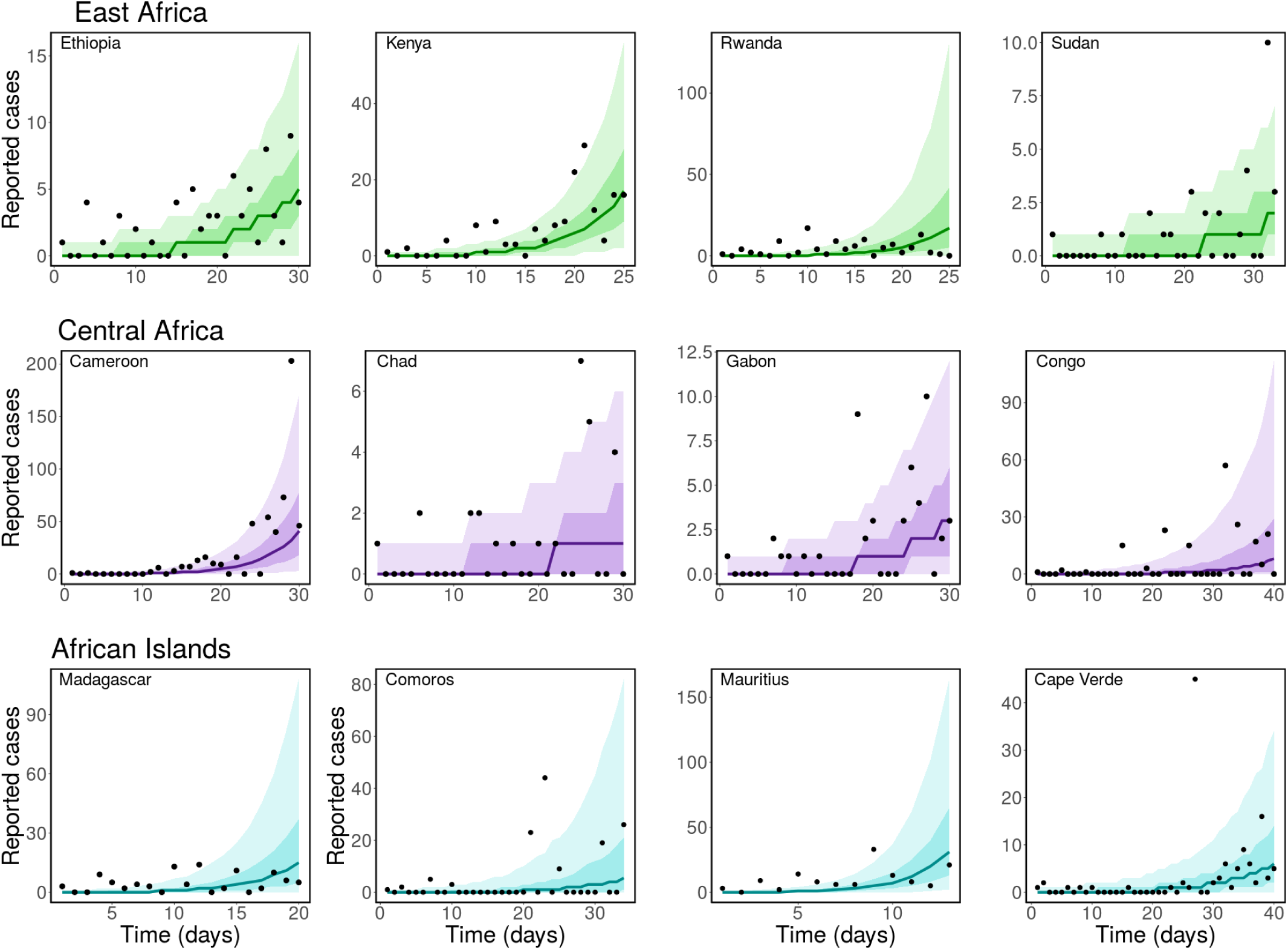
Same caption as Figure 1 but for East African countries: Ethiopia, Kenya, Rwanda and Sudan (top panel), Central African countries: Cameroon, Chad, Gabon and Congo (middle panel), and African Islands: Madagascar, Comoros, Mauritius, and Cape Verde (bottom panel).

**Figure 3:**
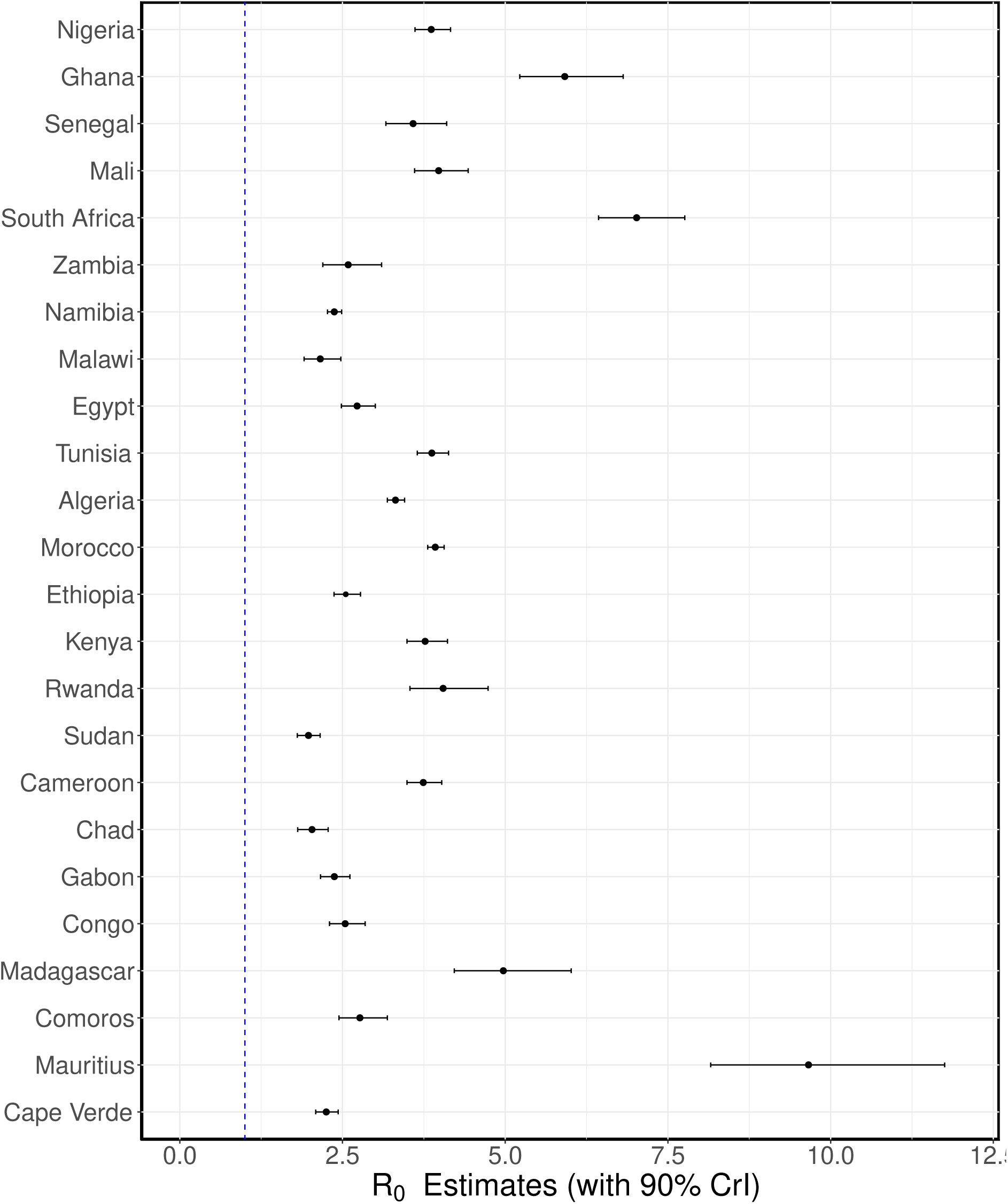
Estimated basic reproduction numbers. The estimated basic reproduction number for each country. The black dots are the median estimates of the reproduction numbers, while thethin bars are 90% credible intervals (CrI).

In Figure 1, we have the fits for the West African countries: Nigeria, Ghana, Senegal and Mali (top panel), Southern African countries: South Africa, Zambia, Namibia and Malawi (middle panel), and the North African countries: Egypt, Tunisia, Algeria and Morocco (bottom panel). We estimated the basic reproduction number for Nigeria to be *R*_0_ = 3.86 (90% CrI : 3.61 − 4.16). The first case of COVID-19 in Nigeria was reported on February 27, 2020. However, the first non-pharmaceutical intervention in Nigeria was implemented 20 days after the index case. Between March 18 - April 10, several non-pharmaceutical interventions (NPIs) ranging from travel restrictions, lockdown, stay-at-home orders, among others, were implemented in some states [27]. The implementation of these NPIs aid the decrease in COVID-19 cases in Nigeria during the early stage of the epidemic. For Ghana, we estimated *R*_0_ = 5.91 (90% CrI : 5.22 − 6.81). Observe that the estimated *R*_0_ for Ghana is larger than that of Nigeria. This is probably due to the rapid increase in cases observed in Ghana in the first 15 days. By day 10, Ghana had already started reporting daily cases in high twenties. Specifically, 26, 40, and 39 cases where reported on day 11, 12 and 13, respectively while the daily reported cases in Nigeria within the first twenty days were all less than 20. Travel restrictions were imposed on March 22, 2020 in Ghana, and lockdown imposed in the Greater Accra Metropolitan Area and the Greater Kumasi Metropolitan Area on March 30 [41]. Even though, there was an initial fast rise in cases in Ghana, the rapid implementation of interventions helped in controlling the spread of the disease. The estimated basic reproduction number for Senegal is *R*_0_ = 3.78 (90% CrI : 3.16 − 4.10), while for Mali we estimated *R*_0_ = 3.98 (90% CrI : 3.61 − 4.43). Mali is one of the countries in Africa that implemented NPIs before their first case of COVID-19. Travel restrictions, closure of schools, and ban on public gathering were imposed on March 18, 2020, while their first case of COVID-19 was reported on the 25^th^ of March, 2020 [4]. The plots of the posterior prediction for these countries are presented in the top panel of Figure 1, while the estimated *R*_0_ values with 90% credible intervals are plotted in Figure 1 and presented in Table 2.

In the middle row of Figure 1, we show the results of our inferences for the Southern African countries. In the left panel of this row, we have the result for South Africa, where we estimated *R*_0_ = 7.02 (90% CrI : 6.43−7.76). Followed by Zambia with *R*_0_ = 2.59 (90% CrI : 2.20−3.10) in the middle-left panel and Namibia with *R*_0_ = 2.37 (90% CrI : 2.27 − 2.49) in the middle-right panel. The result in the right panel of this row is for Malawi, where the basic reproduction number was estimated as *R*_0_ = 2.16 (90% CrI : 1.91 − 2.47). We observe from these results that South Africa has the largest basic reproduction number for this region followed by Zambia, Namibia, and then Malawi. The ordering of the estimated *R*_0_ for these countries also reflect the current situation of the pandemic in the region. As of September 30, 2021, South Africa has the highest reported cases COVID-19 with 2,902,672 cases, followed by Zambia: 209,046 reported case, Namibia: 127,589 cases, and Malawi: 61,580 reported cases [102]. The estimated basic reproduction numbers for these countries are also plotted in Figure 3, and given in Table 2 with 90% credible interval.

The results for North African countries are given in the bottom panel of Figure 1. The largest basic reproduction number for this region was estimated for Morocco, *R*_0_ = 3.92 (90% CrI : 3.81 − 4.06), followed by Tunisia with an estimate of *R*_0_ = 3.87 (90% CrI : 3.65 −4.13), Algeria: *R*_0_ = 3.31 (90% CrI : 3.19 −3.45), and Egypt *R*_0_ = 2.72 (90% CrI : 2.48 − 3.00). As of September 30, 2021, Morocco also has the largest epidemic in this region, followed by Tunisia, Egypt and Algeria [102]. Egypt has the sixth largest COVID-19 epidemic in Africa as of September 30, 2021, even though, it reported the continent’s first case of the disease [102].

Similar results are presented in Figure 2 for East African countries: Ethiopia, Kenya, Rwanda and Sudan (top panel), Central African countries: Cameroon, Chad Gabon and Congo (middle panel), and African island countries/regions: Madagascar, Comoros, Mauritius, and Cape Verde (bottom panel). Sudan has the smallest estimated basic reproduction number for the East African countries considered, with *R*_0_ = 1.98 (90% CrI : 1.81 − 2.16), followed by Ethiopia with *R*_0_ = 2.55 (90% CrI : 2.37 − 2.78), Kenya with *R*_0_ = 3.77 (90% CrI : 3.49 − 4.11), and Rwanda with an estimated *R*_0_ = 4.04 (90% CrI : 3.52 − 4.72). The posterior prediction for these countries are presented in the first row of Figure 2. In the second row of the same figure, we show the results of our inferences for the Central African countries considered. For this region, the estimated basic reproduction numbers are *R*_0_ = 3.74 (90% CrI : 3.49 − 4.02) for Cameroon, *R*_0_ = 2.03 (90% CrI : 1.81 − 2.28) for Chad, *R*_0_ = 2.37 (90% CrI : 2.16 − 2.61) for Gabon, and *R*_0_ = 2.54 (90% CrI : 2.30 − 2.85) for the Republic of the Congo. Among these countries, Cameroon was the first to have a COVID-19 case (Mar 6), followed by the Republic of the Congo (Mar 10), Gabon (Mar 13), and Chad (Mar 19). As of September 30, 2021, there had been 92,303 cases of COVID-19 in Cameroon, 5, 038 in Chad, 30, 155 in Gabon, and 14, 244 cases in Congo. Even though, the basic reproduction number estimated for Chad is larger than that of Congo, we observe that the reported cases of the disease in Congo is almost three folds more than that of Chad as of end September 2021. This may be as a result of different factors ranging from the intervention strategies implemented in these countries and how strict they are imposed, their population sizes, the mixing patterns in the populations, among others. See Table A2 of Appendix A and references therein for more details about the population sizes of these countries and the intervention strategies implemented by the government during the early stage of the disease.

Lastly, we present the results for African island countries in the bottom row of Figure 2. In the left panel on this row, we have the result for Madagascar, where the basic reproduction number is estimated as *R*_0_ = 4.97 (90% CrI : 4.22 − 6.01). For Comoros, with estimated *R*_0_ = 2.77 (90% CrI : 2.45 − 3.19), the posterior prediction is shown in the middle-left panel, for Mauritius, we estimated *R*_0_ = 9.66 (90% CrI : 8.15 − 11.75), and for Cape Verde, the reproduction number is estimated as *R*_0_ = 2.50 (90% CrI : 2.09 − 2.43). These estimates are also plotted in Figure 3 and presented in Table 2 for easy comparison. The African Islands countries considered have a population size of at least 500,000. Among these countries, Mauritius was the first to report a COVID-19 case on Mar 18, 2020, followed by Madagascar and Cape Verde on Mar 19, 2020, and then Comoros on Apr 30, 2020. As of September 30, 2021, Madagascar had recorded 42, 898 cases of COVID-19 followed by Cape Verde with 37, 576 cases, Mauritius with 15, 695, and then Comoros with 4, 141 reported cases. See Table A2 of Appendix A for the population size of each of these countries, when their index cases were reported, and the intervention strategies implemented during the early stage of the disease.

Overall, our estimates of COVID-19 basic reproduction number for the countries considered ranges between 1.98 (for Sudan) to 9.66 (for Mauritius) with an average of 3.68 and a median of 3.67 (90% CrI: 3.31 - 4.12). Since *R*_0_ gives the expected number of secondary cases caused by a single infected individual introduced into a completely susceptible populations, countries that reported high daily COVID-19 cases consistently during the early days of their outbreak tend to have higher basic reproduction numbers. However, this does not necessarily imply that these countries would have larger epidemics as some of them imposed strict non-pharmaceutical interventions, such as lockdown, that helped to control the spread of the disease. Countries with low population sizes and high daily reported cases of COVID-19 during the first few days of their outbreak also tend to have larger estimates of *R*_0_ compared to those with larger population sizes. For most of the countries we considered and in each group, the ordering of their estimated basic reproduction number reflects the ordering of their epidemic sizes as of September 30, 2020. Observe that the fits for the Northern African countries, shown in the bottom panel of Figure 1, look relatively better than those of the other regions. This reflects the quality of the data from these countries and emphasizes the importance of have a consistent data set.

In this paper, we have assumed that the population of each country is well-mixed (homogeneous) and modelled their population as a single population. In reality, the contact rates and mixing pattern between individuals in different regions of the country may vary. As a result, the initial spread of COVID-19 in a country will depend on the contact rate of individuals in the region of the country where the index case occurred. The spread would also depend on the rate people travel out of the region to other parts of the country. Our modeling framework does not explicitly capture the mobility of individuals in the country. Another limitation of this work includes not accounting for under-reporting. Due to the highly contagious nature of SARS-CoV-2, many countries advised individuals with COVID-19-like symptoms to self-isolate at home instead of going to the hospital, except they have a severe case of the disease. As a result, these individuals may not get a test to confirm if they had the disease, and therefore, they may not be captured in the reported cases. This may lead to under-reporting. The availability and consistency of data is also another limitation of this work. Data collection during the COVID-19 pandemic has been very tedious worldwide, especially during the early stage of the disease when non-pharmaceutical interventions that impede movement and access to resources were imposed. Particularly, data collection in many developing countries across Africa is challenging due to the lack of adequate infrastructure, leading to inadequate data. The framework presented in this paper requires knowing the transition rates of infected individuals between the different compartments of the model. These parameters may not be immediately available for a novel disease like SARS-CoV-2 during the early stage of the disease. Another limitation of our model includes not explicitly modeling the dynamics of asymptomatic individuals.

An interesting extension of the work presented in this paper includes incorporating a parameter for the ascertainment fraction of the reported cases of COVID-19 in each countries. In this case, the ascertainment fraction can be estimated together with the basic reproduction number using Bayesian inference. We know that the reported cases of COVID-19 around the world does not capture all the cases of the disease. It would be worthwhile to infer the actual cases of the disease in each country. Another extension of this work is to incorporate a compartment that explicitly account for the population of asymptomatic individuals in each country. In this case, infected individuals transitioning from the exposed to infectious compartment can either become symptomatic or asymptomatic.

## Conclusion

The COVID-19 disease continues to spread all over the world despite the availability and deployment of vaccines. As a result, there is still the need to adequately understand the transmission dynamics of the disease. In this study, we used a simple susceptible-exposed-infectious-recovered (SEIR) model together with a Bayesian inference framework to estimate the basic reproduction number of COVID-19 in selected African countries. These estimates are based on the reported number of COVID-19 cases in these countries during the early stages of their epidemic before major non-pharmaceutical interventions were implemented. The results obtained in this study will help inform mathematical models used to study the dynamics of COVID-19 in the selected African countries. In addition, the *R*_0_ estimates can be used by public health officers and government officials to adequately plan and map out strategies that will help eradicate the disease.

## Data Availability

All data produced in the present work are contained in the manuscript

## Data Availability

All the cases data used in this research are public available on the GitHub page of *Our World in Data:*

https://github.com/owid/covid-19-data/blob/master/public/data/README.md.

## A Appendix

**Table A1:**
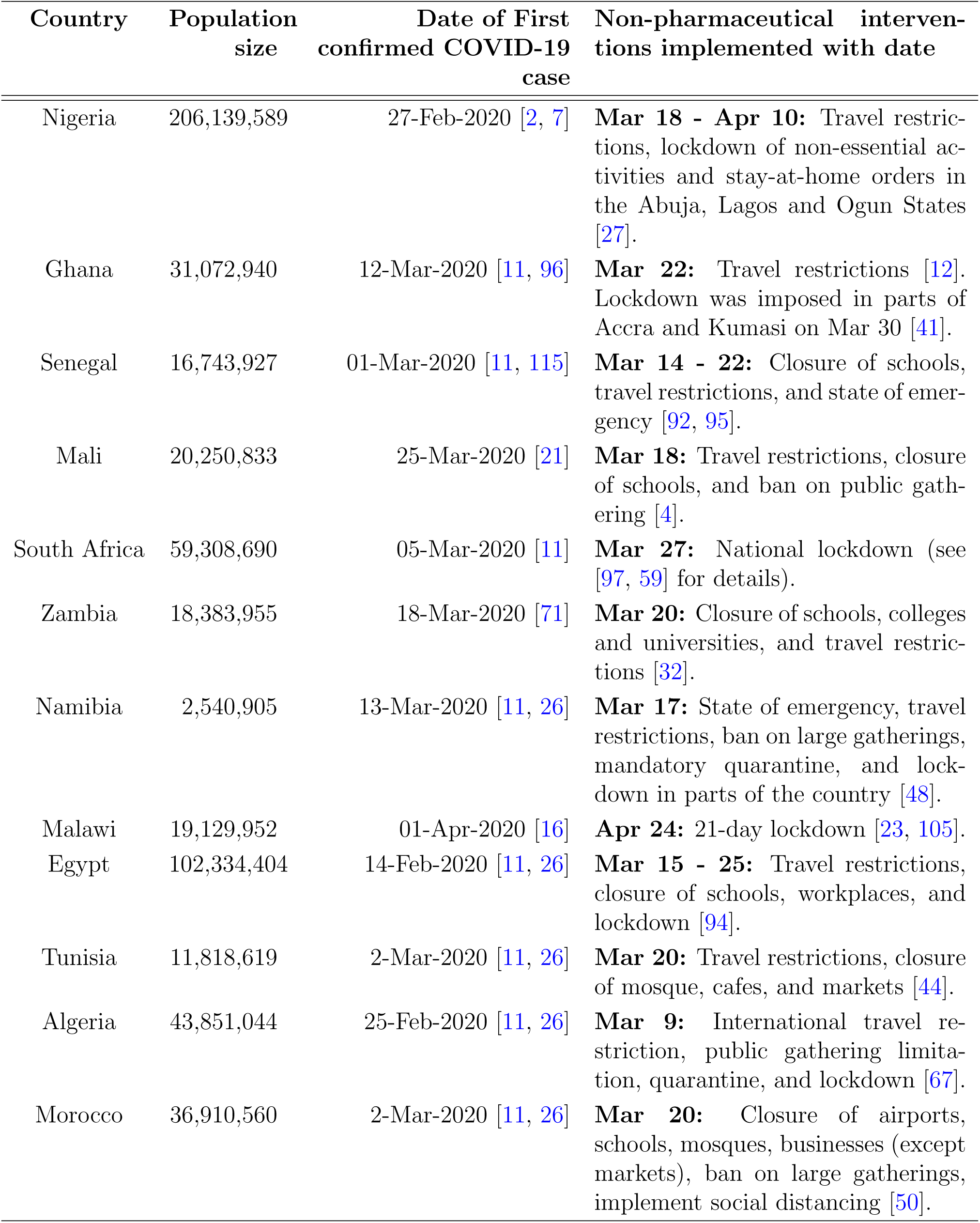
List of countries, their population sizes, date of their index case of COVID-19, and the non-pharmaceutical interventions (NPIs) implemented against COVID-19 in the early stage of the disease in the countries with dates. The population sizes are for the year 2020 and were obtained from the worldometer population website [116]. When an interval is specified for the date of NPIs, it means the implementation of the NPIs started on a day within the specified interval.

**Table A2:** Same caption as Table A1

| Country | Population size | Date of First confirmed COVID-19 case | Non-pharmaceutical interventions implemented with date |
| --- | --- | --- | --- |
| Ethiopia | 114,963,588 | 13-Mar-2020 [11, 26] | **Mar 16 - 23:** Suspension of schools, sporting events, and public gatherings for 15 days. Closure of night clubs and all land borders [3, 20]. |
| Kenya | 53,771,296 | 13-Mar-2020 [11, 26] | **Mar 15-25:** Closure of all learning institutions, bars and restaurants, ban on social gathering, travel restrictions, dusk to dawn curfew imposed [56] |
| Rwanda | 12,952,218 | 14 March 2020 [77] | **Mar 20:** National lockdown and banning of all non-essential travel, labour, and businesses [51]. |
| Sudan | 43,849,260 | 13-Mar-2020 [11, 26] | **Mar 16 - 24:** Closure of airports, nationwide curfew and lockdown [72]. |
| Cameroon | 26,545,863 | 6-Mar-2020 [26] | **Mar 18:** Closure of land, air and sea borders and mandatory mask wearing in public spaces on April 13 [80]. |
| Chad | 16,425,864 | 19-Mar-2020 [106] | **Apr 13:** Closure of schools and workplaces, and partial lockdown in different regions [93]. |
| Gabon | 2,225,734 | 13-Mar-2020 [11, 26] | **Jul 1:** Suspension of visas for all European travelers [91]. |
| Republic of the Congo | 5,518,087 | 10-Mar-2020 [26] | **Mar 31:** Nationwide lockdown [79]. |
| Madagascar | 27,691,018 | 19-Mar-2020 [90] | **Mar 20 - 31:** Suspension of international and regional flights [42]. Lockdown in at least two cities [69]. |
| Comoros | 869,601 | 30-Apr-2020 [43] | **Apr 30:** Curfew imposed, and mandatory wearing of masks outdoors on the whole territory implement on July 8 [39, 38] |
| Mauritius | 4,649,658 | 18-Mar-2020[24] | **Mar 20:** Nationwide "sanitary" lockdown. Essential services and certain economic services, and minimum public transport are allowed to operate (see [29] for more details). |
| Cape Verde | 555,987 | 19-Mar-2020 [46] | **Mar 28:** Nationwide state of emergency imposed [28]. |

